# Depression, anxiety, and chronic fatigue symptoms in acute rheumatoid arthritis are associated with immune-inflammatory, autoimmune, endogenous opioid system and lactosylceramide signaling pathways: a nomothetic network approach

**DOI:** 10.1101/2021.09.26.21264149

**Authors:** Hasan Najah Smesam, Hasan Abbas Qazmooz, Sinan Qayes Khayoon, Hussein Kadhem Al-Hakeim, Michael Maes

**Author notes:** Corresponding author: Prof. Dr. Michael Maes, M.D., Ph.D., IMPACT Strategic Research Center, Barwon Health, Deakin University, Geelong, Vic, Australia. https://scholar.google.co.th/citations?user=1wzMZ7UAAAAJ&hl=th&oi=ao.

## Abstract

**Background:** Rheumatoid arthritis (RA) is a chronic inflammatory and autoimmune disorder which affects the joints in the wrists, fingers, and knees. RA is often associated with depressive and anxiety symptoms as well as chronic fatigue syndrome (CFS)-like symptoms.

**Aim:** To examine the association between depressive symptoms (measured with the Beck Depression Inventory, BDI), anxiety (Hamilton Anxiety Rating Scale, HAMA), and CFS-like (Fibro-fatigue Scale) symptoms and immune-inflammatory, autoimmune, and endogenous opioid system (EOS) markers, and lactosylceramide in RA.

**Methods:** Serum biomarkers were assayed in RA patients with (n=59) and without (n=59) increased psychopathology (PP) and 50 healthy controls.

**Results:** There were highly significant correlations between the BDI, FF, and HAMA scores and severity of RA, as assessed with the DAS28-4, clinical and disease activity indices, the number of tenders and swollen joints, and patient and evaluator global assessment scores. A common latent vector (reflective model) could be extracted from the PP and RA-severity scales, which showed excellent psychometric properties. Partial least squares analysis showed that 69.7% of the variance in this common core underpinning PP and RA symptoms could be explained by the regression on immune-inflammatory pathways, rheumatoid factor and anti-citrullinated protein antibodies, CD17, and mu-opioid receptor levels.

**Conclusions:** Depression, anxiety, and CFS-like symptoms due to RA are reflective manifestations of the phenome of RA and are mediated via the effects of the same immune-inflammatory, autoimmune, and EOS pathways and lactosylceramide that underpin the pathophysiology of RA. These PP symptoms are clinical manifestations of the pathophysiology of RA.

## Introduction

Rheumatoid arthritis (RA), a chronic autoimmune disease (CAID) that causes progressive damage to bone, cartilage, and joints in the hands and feet, may result in synovial membrane thickening and deformation and disabilities. ^1–3^ Inflammation of the synovial joints, which leads to joint pulverization and destruction, is associated with autoimmune responses to rheumatoid factor (RF) and anti-cyclic citrullinated peptide (anti-CCP), increased immune complexes, activated immune-inflammatory and nitro-oxidative pathways. ^4, 5^ The inflammatory mediators that cause joint inflammation, including the proinflammatory cytokines interleukin (IL)-1β, IL-6, and tumor necrosis factor (TNF)-α, may spread systemically, leading to comorbid disease, including cardiovascular, pulmonary, kidney, and gastrointestinal disease. ^6, 7^ IL-6 plays a significant role in the development of RA and is a key inducer of the acute phase response, resulting in elevated levels of C-Reactive Protein (CRP). ^8, 9^ IL-6 is synthesized by endothelial cells and lymphocytes and synthesizes acute reactive proteins that can damage articular cartilage. ^10, 11^ The granulocyte-macrophage colony-stimulating factor (GM-CSF) is an important factor promoting monocyte/macrophage survival in synovial liquid and membrane. ^12, 13^ GM-CSF may aggravate arthritis in RA animal models, while a lack of functional GM-CSF in collagen-induced arthritis in mice shows protective activities. ^13, 14^ IL-10, a negative immune regulatory cytokine, is often elevated in RA and is linked to increased RF and anti-CCP antibodies. ^15^

Some studies have suggested that inflammation and joint-destructive pathways in collagen-induced arthritis may be mediated by the Toll-Like-Receptor (TLR)4 and that TLR4 deficiency is associated with reduced autoimmune responses as indicated by lower IL-17 and anti-CCP levels. ^16, 17^ TLRs are abundant in synovial tissues from RA patients, and TLR ligands may activate synovial fibroblasts and immune cell chemotaxis. ^18^

Some ceramides (lipid second messengers) mediate IL-1β and TNF-α signaling in RA synovial cells and cause apoptosis. ^19^ One of the ceramides, lactosylceramide (CD17), is involved in chemotaxis, phagocytosis, and superoxide generation, ^20, 21^ as well as activating NADPH oxidase, resulting in the production of superoxide radicals (O^2^.) ^22^ and inducible nitric oxide synthase (iNOS). ^23^ There is also a report that RA is associated with changes in the endogenous opioid system (EOS), as evidenced by an inverse correlation between plasma β-endorphin levels, rheumatoid disease activity scores, and RA duration. ^24^ Acute inflammation with increased TNF-α and IL-1β levels causes leukocytes to release β-endorphins and mu (MOR) and kappa (KOR) opioid receptors. ^25, 26^ These EOS compounds have antinociceptive and anti-inflammatory properties ^26^, suggesting that they may play a role in the pathophysiology of RA.

A significant proportion of RA patients exhibit symptoms of depression ^27, 28^, anxiety, ^29–31^ and chronic fatigue syndrome (CFS)-like symptoms. ^32, 33^ According to one study, 67.5 percent of RA patients suffer from depression, with 60 percent suffering from moderate to severe depression. ^34^ CFS-like symptoms can be found in 40-70 percent of RA patients, with 41 percent experiencing severe fatigue. ^35^ The activated immune-inflammatory and nitro-oxidative pathways in that disorder may explain this high incidence of affective and CFS-like symptoms in RA. ^36^ Affective disorders including major depressive disorder (MDD) and generalized anxiety disorder, and CFS are neuro-immune and neuro-oxidative disorders characterized by immune activation (increased IFN-γ and IL-10), chronic low-grade inflammation (increased IL-1β, IL-6, and TNF-α), and increased nitro-oxidative stress (lipid peroxidation, protein oxidation). ^37–43^ In RA, depressive symptoms are significantly associated with disease activity and immune activation markers, such as increased levels of TNF-α, IL-2, IL-4, and IL-6. ^44^ Inflammatory symptoms such as elevated CRP levels and tender and swollen joints are linked to increased fatigue. ^45^ A review reported that RA’s baseline IL-1 and interferon (IFN)-γ levels predict increased fatigue several weeks later. ^46^

TLR4 complex activation is thought to be important in affective disorders and CFS. ^47^ Ceramides, including lactosylceramide (CD17), are significantly higher in depressed patients than in controls. ^48–50^ Increased levels of CD17 are significantly associated with the severity of affective symptoms in people with type 2 diabetes mellitus (T2DM). ^51^ Increased levels of β-endorphin and MOR have been found in major depression, and it is thought that they act as part of the negative immune-regulatory system (CIRS). ^52–54^ Furthermore, we recently discovered an inverse relationship between the F-box/WD repeat-containing protein 7 (FBXW7) and affective symptoms, which was mediated by increased atherogenicity and insulin resistance. ^51^ FBXW7 is an E3 ubiquitin ligase that catalyzes the ubiquitin-mediated degradation of cyclin E ^55^. FBXW7 controls neurogenesis by antagonizing c-Jun and Notch, two transcription factors required for neuronal differentiation and synaptic plasticity. ^56^

However, the precise relationships between affective and CFS-like symptoms due to RA and the above-mentioned biomarkers and severity of RA are not well defined. Hence, the purpose of this study was to examine if affective and CFS-like symptoms due to RA are associated with RA severity, autoimmune biomarkers (RF and anti-CCP), signs of immune activation (IL-6, CRP, GM-CSF, IL-10, soluble TLR4), and other biomarkers including CD17, FBXW7, and EOS biomarkers (β-endorphins, MOR, KOR, and endomorphin-2). The specific hypothesis is that affective and CFS symptoms due to RA are associated with increased immune-inflammatory biomarkers (IL-6, CRP, GM-CSF, TLR4, IL-10), increased RF and anti-CCP antibody levels, CD17 and EOS compounds (KOR, MOR, and β-endorphin) but lowered FBXW7.

## Materials and Methods

### Participants

This case-control study was conducted in Al-Sader Teaching Hospital in Al-Najaf province-Iraq, from February 2021 till June 2021. The current study included 118 RA patients as well as 50 age- and gender-matched healthy controls. The “American College of Rheumatology” and the “European League Against Rheumatism” arthritis criteria were used to diagnose definite RA ^57^ based on synovitis in at least one joint and a score of 6 or more from scores in 4 domains, namely numbers and sites of affected joints, serologic abnormalities (RF and anti-CCP antibodies), acute-phase response (CRP), and symptom duration. Healthy controls were recruited from the same catchment area as family members or friends of staff and friends of patients. They were age and sex-matched with the patients. Exclusion criteria for patients and controls were the presence of any other systemic disease, including liver and heart disease and diabetes, other axis-1 psychiatric disorders, and neuroinflammatory or neurodegenerative disorders (including stroke, multiple sclerosis, Parkinson’s disorder, etc.). In addition, controls were excluded when they suffered from CFS, CF, Myalgic Encephalomyelitis, and RA or rheumatoid arthritis-like symptoms and showed a lifetime diagnosis of any axis-1 psychiatric disorder, including major depression and bipolar disorder, and dysthymia.

The Disease Activity Score (DAS28-CRP) was calculated using the calculator (https://www.das-score.nl/das28/DAScalculators/dasculators.html) based on the tender joint count (TJC), swollen joint count (SJC), CRP, and the patient’s general health measured using a visual analog scale. The online calculator https://www.mdcalc.com/clinical-disease-activity-index-cdai-rheumatoid-arthritis was employed to calculate the Clinical Disease Activity Index (CDAI). The simple disease activity index (SDAI) was computed using the online calculator https://www.mdcalc.com/simple-disease-activity-index-sdai-rheumatoid-arthritis. The patient global assessment (PGA) score was a patient-reported outcome which was assessed as a single question scored from 0-10 (“how is your health overall”). ^58^ The evaluator global assessment (EGA) assessed indexed disease activity (0-10 score) was evaluated by a senior rheumatologist. ^59^ The same day as the assessment of the severity of RA, a senior psychiatrist assessed the severity of depression using the Beck-Depression Inventory (BDI-II) score, ^60^ anxiety using the Hamilton Anxiety Rating Scale (HAMA), ^61^ and Fibromyalgia and Chronic Fatigue Syndrome Rating (FF) scale to measure the severity of CFS- and fibromyalgia-like symptoms. ^62^ An overall index of severity of psychopathology was computed as z total BDI-II score + z total FF score + z total HAMA score (z PP score). Using the median of the z PP score, we dichotomized the patient groups into two subgroups, namely patients with (RA+PP) and without (RA) increased z PP. We also computed two BDI-II subdomain scores, namely a) key depressive (Key_BDI) symptoms (sum of the items sadness, pessimism, loss of pleasure, loss of interest, past failure, guilty feelings, punishment feelings, self-dislike, self-criticalness, worthlessness), and 2) physiosomatic BDI-II (PS_BDI) symptoms (sum of the items loss of energy, changes in sleeping pattern, changes in appetite, concentration difficulties, tiredness or fatigue, loss of interest in sex). We also computed two HAMA subdomain scores, namely a) key anxiety (Key_HAMA) symptoms, namely sum of the HAMA items, anxious mood, tension, fears, and anxiety behavior at interview; and 2) physiosomatic HAMA (PS_HAMA) symptoms, namely sum of 6 symptoms, i.e., somatic sensory, and cardiovascular, respiratory, gastrointestinal, genitourinary and autonomic symptoms. Key FF (Key_FF) symptoms were computed as the sum of 5 symptoms: fatigue, autonomous and gastrointestinal symptoms, headache, and a flu-like malaise (thus excluding the cognitive, affective, and rheumatoid-like symptoms). The total physiosomatic score (z PS) was the sum of z PS_BDI score + z PS_HAMA + z key FF. We also computed a composite score reflecting “chronic fatigue” (z fatigue) as z BDII fatigue + z FF fatigue. Body mass index (BMI) was assessed the same day as the clinical interview as body weight in kg/length in m^2^.

The ethics committee (IRB) of the University of Kufa, Iraq (243/2020), which complies with the International Guideline for Human Research Protection, approved the study as required by the Helsinki Declaration. Written informed consent was obtained from patients and controls before inclusion in the study.

### Assays

After an overnight fast, all individuals had five milliliters of venous blood drawn and centrifuged for 15 minutes at 3000 rpm after full clotting at 37°C. Sera were collected and kept at −80°C until further analysis. Serum CRP was measured by lateral flow immunoassay kit supplied by Shenzhen Lifotronic Technology^®^ Co. Ltd., China. RF was measured using semi-quantitative kits provided by the Spinreact^®^ Co., Girona, Spain, based on the latex agglutination approach. A semi-quantitative anti-CCP assay was carried out using kits provided by Hotgen Biotech Co., Ltd., Beijing, China.

Commercial ELISA kits were used to measure serum β-endorphin, IL-6, CD17, and FBXW7 (Melsin^®^ Medical Co, Jilin, China), MOR, KOR, EM-2, and GM-CSF (MyBioSource^®^ Inc. San Diego, California, USA), and IL-10 (Elabscience^®^, Hubei, China). The sensitivities of the kits were for IL-6 and β-endorphin: 0.1 pg/mL, IL-10: 4.69 pg/mL: and MOR: 7.18 pg/mL, KOR: 1.0 ng/mL, EM-2: 0.33 pg/mL, CD17 and FBXW7: 0.1 ng/ml, and GM-CSF: 1.0 pg/ml. The intra-assay coefficients of variation (CV%) of all assays were less than 10%. The processes were carried out precisely as prescribed by the manufacturer, with no deviations. Sera were diluted at 1:4 to estimate CD17 levels.

### Statistical analysis

The Chi-square test was used to assess associations between categorical variables, and the analysis of variance (ANOVA) test was used to compare scale variables across categories. We used Pearson’s product-moment correlation analysis and the point-biserial correlation analysis (the latter to assess the relationships between dichotomous variables (RF and anti-CCP) and continuous variables (e.g., clinical data). Multivariate generalized linear model (GLM) analysis was utilized to check associations between diagnostic classes (RA subgroups and controls) and the measured biomarkers while controlling for age, sex, smoking and BMI entered as covariates. Subsequently, we used between-subject effects to check the associations between diagnosis and each significant biomarker. In this investigation, the partial eta-squared (η^2^) effect size was utilized. Automatic multiple regression analysis was used to predict dependent variables (clinical rating scale scores) based on biomarkers and demographic data while checking for R^2^ changes, multivariate normality (Cook’s distance and leverage), multicollinearity (using tolerance and VIF), and homoscedasticity (using White and modified Breusch-Pagan tests for homoscedasticity). We used an automatic stepwise (step-up) method with 0.05 p-to-enter and 0.06 p-to-remove. The results of these regression analyses were always bootstrapped with 5.000 bootstrap samples, and the latter results are displayed if the results were not concordant. All tests are two-tailed, and a p-value of 0.05 was used to determine statistical significance. For statistical analysis, IBM SPSS Windows version 25, 2017, was used.

The SmartPLS path analysis method was used to determine the causal relationship between biomarkers and the RA phenome. Each variable was entered as either a single indicator or a latent vector derived from its reflective manifestations. When the outer and inner models complied with pre-specified quality data, complete PLS analysis was performed using 5000 bootstrap samples, namely a) the model fit SRMR is < 0.08, b) all latent vectors have good composite reliability (> 0.7), Cronbach’s alpha (> 0.7), and rho A (> 0.8), with an average variance extracted > 0.5, c) all loadings on the latent vectors are > 0.666 at; and d) the latent vectors were not misspecified as reflective models (tested with Confirmatory Tetrad Analysis, CTA). Complete PLS analysis was carried out with 5.000 bootstrap samples and specific indirect, total indirect, and direct pathway coefficients (with exact p-value) were computed. The predictive performance was evaluated using PLS predict with 10-fold cross-validation Predicted-Oriented Segmentation analysis, Multi-Group Analysis, and Measurement Invariance Assessment were used to investigate compositional invariance. ^63^

## Results

### Sociodemographic data

**Table 1** shows the sociodemographic and clinical data as well as PP rating scale scores in RA patients with RA+PP versus RA and the control group. We found no differences in sex, age, and BMI among the three groups. The RA+PP group showed higher CDAI and SDAI scores and TJCs and SJCs than the RA and control groups. The DAS28-4, total BDI-II. HAMA and FF scores increased from controls → RA → RA+PP group.

**Table 1.** Sociodemographic and clinical data in healthy controls (HC) and patients with rheumatoid arthritis divided into those with (RA+PP) and without (RA) increased psychopathology

| Variables | HC <sup>A</sup><br>n=50 | RA <sup>B</sup><br>n=59 | RA+PP <sup>C</sup><br>n=59 | F/$\chi^2$ | df | p |
| --- | --- | --- | --- | --- | --- | --- |
| Age years | 48.7 ±4.6 | 49.8 ±7.3 | 50.3 ±6.8 | 0.90 | 2/165 | 0.407 |
| Sex F/M | 27/23 | 36/23 | 33/26 | 0.60 | 2 | 0.471 |
| BMI kg/m<sup>2</sup> | 27.44 ±2.43 | 27.85 ±2.65 | 27.60 ±2.87 | 0.33 | 2/165 | 0.721 |
| TUD No/Yes | 44/6 <sup>C</sup> | 57/2 <sup>C</sup> | 45/14 <sup>A, B</sup> | 10.80 | 2 | 0.005 |
| RF No/Yes | 50/0 <sup>B, C</sup> | 14/45 <sup>A, C</sup> | 9/50 <sup>A, B</sup> | 93.50 | 2 | <0.001 |
| ACPA No/Yes | 50/0 <sup>B, C</sup> | 27/32 <sup>A, C</sup> | 24/35 <sup>A, B</sup> | 47.54 | 2 | <0.001 |
| DAS28-4 | 1.61 ±0 <sup>B, C</sup> | 4.50 ±0.65 <sup>A, C</sup> | 5.26 ±0.67 <sup>A, B</sup> | 650.49 | 2/165 | <0.001 |
| CDAI | 0.0 | 16.7 ±6.0 <sup>C</sup> | 22.9 ±7.1 <sup>B</sup> | KWT | - | <0.001 |
| SDAI | 0.0 | 20.1 ±6.7 <sup>C</sup> | 28.3 ±7.7 <sup>B</sup> | KWT | - | <0.001 |
| PGA | 0.0 | 4.99 (1.97) <sup>C</sup> | 6.31 (2.23) <sup>B</sup> | KWT | - | <0.001 |
| EGA | 0.0 | 4.36 (1.89) <sup>C</sup> | 5.72 (2.15) <sup>B</sup> | KWT | - | <0.001 |
| Tender joint count | 0.0 | 4.9 ±2.2 <sup>C</sup> | 7.2 ±2.7 <sup>B</sup> | KWT | - | <0.001 |
| Swollen joint count | 0.0 | 2.4 ±1.6 <sup>C</sup> | 3.7 ±1.4 <sup>B</sup> | KWT | - | <0.001 |
| Total BDI-II | 8.9 ±2.0 <sup>B, C</sup> | 11.3 ±5.9 <sup>A, C</sup> | 27.5 ±11.0 <sup>A, B</sup> | 103.62 | 2/165 | <0.001 |
| Total FF | 5.9 ±1.7 <sup>B, C</sup> | 12.8 ±5.3 <sup>A, C</sup> | 24.5 ±6.2 <sup>A, B</sup> | 203.44 | 2/165 | <0.001 |
| Total HAMA | 6.0 ±1.7 <sup>B, C</sup> | 13.2 ±3.9 <sup>A, C</sup> | 23.1 ±4.7 <sup>A, B</sup> | 288.14 | 2/165 | <0.001 |

<sup>A, B, C</sup>: Pairwise comparison among group means, KWT: Kruskal-Wallis test.
BMI: Body mass index, TUD: Tobacco use disorder, RF: Rheumatoid factor, ACPA: anti-citrullinated protein antibodies, DAS28-4(CRP): Disease Activity Score by four factors including CRP, CDAI: Clinical Disease Activity Index, SDAI: Simple Disease Activity Index, BDI: Beck Depression Inventory, FF: Fibro Fatigue Rating Scale, HAMA: Hamilton Anxiety Rating Scale, PGA: patient's global assessment, and EGA: evaluator's global assessment.

### Multivariate GLM analysis

**Table 2** shows the associations between the diagnostic groups and all biomarkers combined in a multivariate GLM analysis while adjusting for age, sex, BMI, and smoking. A highly significant effect size of diagnosis (0.466) and a moderate effect size of TUD (0.295) were found. Sex, age, and BMI did not significantly affect the biomarker levels. Tests for between-subjects effects showed that diagnosis was highly significantly associated with CRP, IL-10, GM-CSF, IL-6, and TLR4. The association with CD17, KOR, FBXW7, and EP2 showed lower effects sizes. The model-generated estimated marginal means for the eleven biomarkers adjusted for age, BMI, sex, and smoking are shown in **Table 3**. The GM-CSF and CRP levels increased from control → RA → RA+PP. TLR4, FBXW7, CD17, IL-6, IL-10, EP2, and KOR were significantly higher in RA patients than controls. There were no significant differences in β-endorphins and MOR between the study groups.

**Table 2.** Results of multivariate GLM analysis showing the associations between biomarkers and diagnosis while adjusting for background variables.

| Type | Dependent variables | Explanatory variables | F | df | p | Partial $\eta^2$ |
| --- | --- | --- | --- | --- | --- | --- |
| Multivariate | CRP, GM-CSF, TLR4, FBXW7, CD17, IL-6, IL-10, ENDO, MOR, EP2, KOR | HC/RA/RA+PP | 11.50 | 22/290 | <0.001 | 0.466 |
|  |  | Sex | 0.68 | 11/144 | 0.753 | 0.050 |
|  |  | Age | 1.08 | 11/144 | 0.378 | 0.077 |
|  |  | BMI | 0.69 | 11/144 | 0.749 | 0.050 |
|  |  | TUD | 5.47 | 11/144 | <0.001 | 0.295 |
| Tests for between-subject effects | CRP | HC/RA/RA+PP | 234.54 | 2/154 | <0.001 | 0.753 |
|  | IL-10 | HC/RA/RA+PP | 55.45 | 2/154 | <0.001 | 0.419 |
|  | GM-CSF | HC/RA/RA+PP | 33.49 | 2/154 | <0.001 | 0.303 |
|  | IL-6 | HC/RA/RA+PP | 31.96 | 2/154 | <0.001 | 0.293 |
|  | TLR4 | HC/RA/RA+PP | 21.19 | 2/154 | <0.001 | 0.216 |
|  | CD17 | HC/RA/RA+PP | 7.71 | 2/154 | 0.001 | 0.091 |
|  | KOR | HC/RA/RA+PP | 6.56 | 2/154 | 0.002 | 0.079 |
|  | FBXW7 | HC/RA/RA+PP | 6.16 | 2/154 | 0.003 | 0.074 |
|  | EP2 | HC/RA/RA+PP | 5.66 | 2/154 | 0.004 | 0.069 |
|  | MOR | HC/RA/RA+PP | 1.07 | 2/154 | 0.346 | 0.014 |
|  | B-EN | HC/RA/RA+PP | 0.67 | 2/154 | 0.515 | 0.009 |

HC/RA/RA+PP: healthy controls (HC) and patients with rheumatoid arthritis divided into those with (RA+PP) and those without (RA) increased psychopathology. CRP: C-reactive protein, GM-CSF: granulocyte-macrophage colony-stimulating factor, TLR4: Toll-like receptor 4, FBXW7: F-Box and WD Repeat Domain Containing 7, CD17: lactosylceramide, IL: interleukin, B-EN: β-endorphin, EP2: endomorphin-2, KOR: kappa opioid receptor, MOR: mu opioid receptor.

**Table 3.**
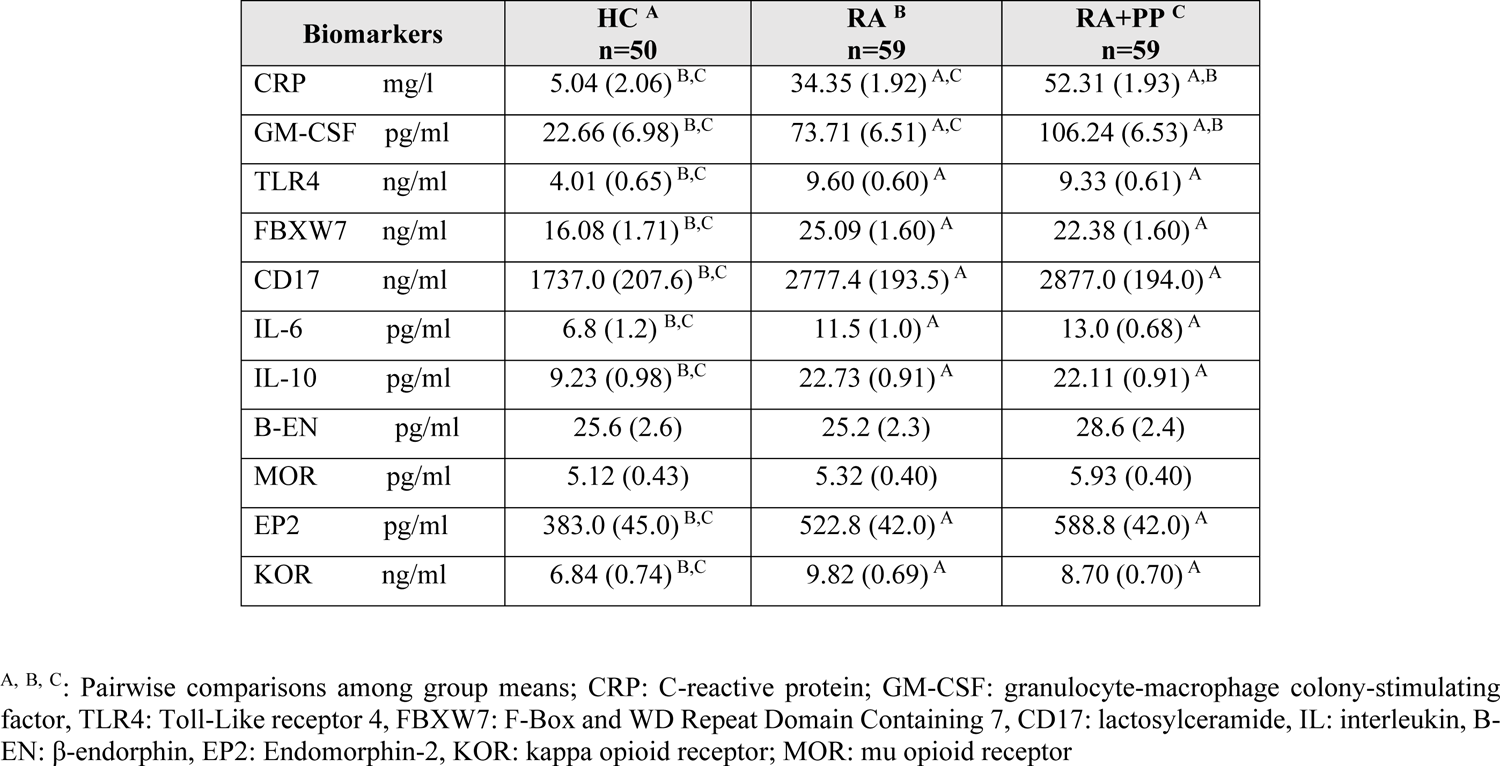
Model-generated estimated marginal means values (SE) of the biomarkers in healthy controls (HC) and patients with rheumatoid arthritis (RA) divided into those with (RA+PP) and without (RA) increased psychopathology scores

### Intercorrelation matrix between psychiatric rating scales and biomarkers

The results of the intercorrelation matrix between psychiatric rating scales and biomarkers are presented in **Table 4**. There is a strong significant correlation between the total BDI-II score and all biomarkers (except FBXW7). The total FF and z PP scores were significantly correlated with all biomarkers. The total HAMA score was significantly associated with all biomarkers except β-endorphins (results of Pearson’s product-moment correlation without p-correction). We have re-run the analysis in the restricted group of RA patients only and found that the correlations between all 4 rating scales and CRP and GM-CSF were significant (all at p<0.01). BDI was associated with TLR4 and IL-6 (at p<0.05). MOR levels were positively associated with FF, HAMA, and z PP values (p<0.05). EP2 was significantly correlated with BDI, FF, and z PP (p<0.01).

**Table 4.** Intercorrelation matrix between psychopathology rating scales and biomarkers.

| **Biomarkers** | **Total BDI** | **Total FF** | **Total HAMA** | **z PP** |
| --- | --- | --- | --- | --- |
| CRP | 0.562*** | 0.834*** | 0.736*** | 0.812*** |
| GM-CSF | 0.408*** | 0.616*** | 0.614*** | 0.634*** |
| TLR4 | 0.304*** / | 0.421*** | 0.435*** | 0.449*** |
| FBXW7 | 0.113 | 0.163* | 0.211** | 0.188* |
| CD17 | 0.244** | 0.286*** | 0.321*** | 0.325*** |
| IL-6 | 0.359*** | 0.471*** | 0.470*** | 0.496*** |
| IL-10 | 0.342*** | 0.554*** | 0.559*** | 0.564*** |
| B-EN | 0.165* | 0.181* | 0.140 | 0.185* |
| MOR | 0.177* | 0.182* | 0.173* | 0.188* |
| EP2 | 0.293*** | 0.339*** | 0.243** | 0.324*** |
| KOR | 0.188* | 0.252** | 0.270*** | 0.275*** |

\*p<0.05; \*\*p<0.01, \*\*\*p<0.001 (n=168).
BDI: Beck's Depression Inventory, FF: Fibro Fatigue scale, Hamilton Anxiety Rating Scale (HAMA), z PP composite score reflecting overall psychopathology
CRP: C-reactive protein; GM-CSF: granulocyte-macrophage colony-stimulating factor; TLR4: Toll-like receptor 4; FBXW7: F-Box and WD Repeat Domain Containing 7; CD17: lactosylceramide; IL: interleukin; B-EN: Beta-endorphin; EP2: Endomorphin-2; KOR: kappa opioid receptor; MOR: mu opioid receptor

### Prediction of the psychopathology scores using biomarkers

**Table 5** shows the results of multiple regression analyses with the PP scale scores as dependent variables and biomarkers as explanatory variables while allowing for the effects of age, sex, smoking, and BMI. Regression #1 shows that 57.9% of the variance in the z PP score was explained by the regression on CRP, GM-CSF, and TLR4. **Figure 1** shows the partial regression plot of the z PP composite score on CRP (adjusted for age, sex, BMI, and IL-10). Regression #2 shows that 60.3% of the variance in the total BDI-II score was explained by CRP, GM-CSF, and TLR4. A considerable part of the variance in the total FF score (60.3%) was explained by CRP, GM-CSF, and EP2 (Regression #3). Regression #4 shows that 58.8% of the variance in the total HAMA score could be explained by the combined effects of CRP, GM-CSF, RF, anti-CCP, and TLR4. Regression #5 shows that 28.1% of the variance in Key_BDI was explained by the combined effects of CRP and GM-CSF. Regression #6 shows that 41.0% of the variance in the Key_HAMA symptom severity was explained by the combined effects of CRP, GM-CSF, and RF.

**Figure 1.**
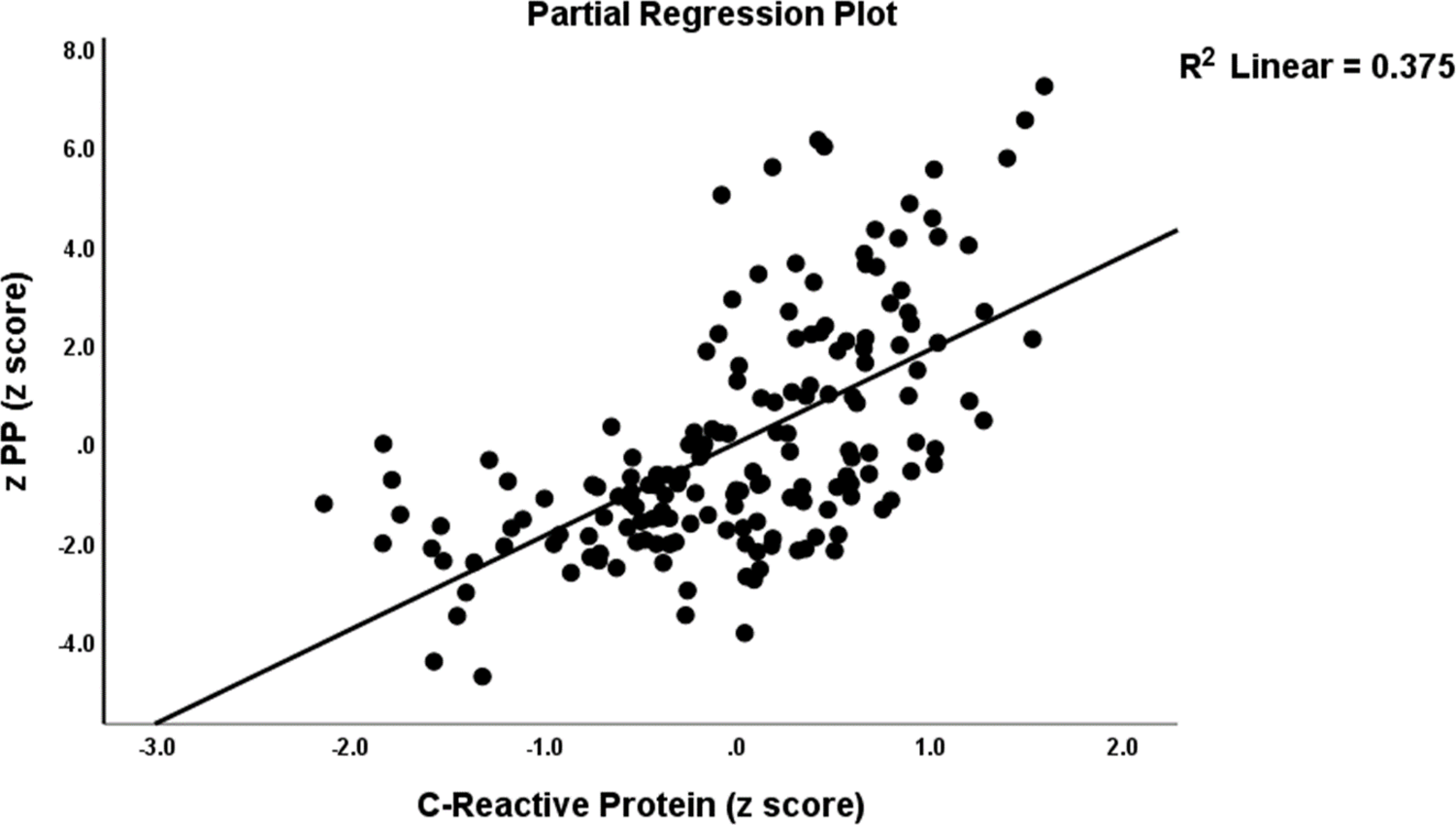
Partial regression plot of the integrated index of psychopathology (z PP) on C-Reactive Protein.

**Table 5.**
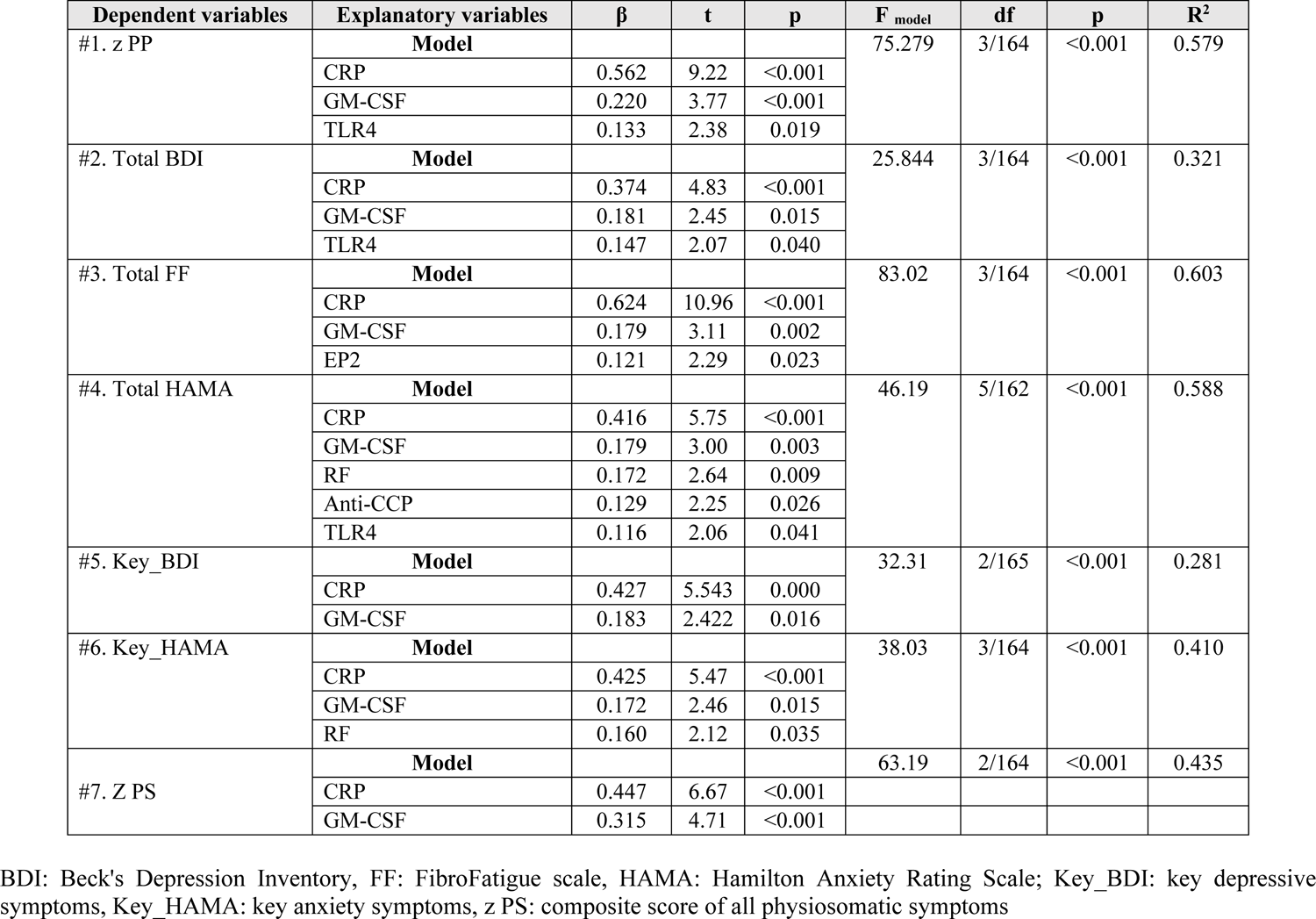
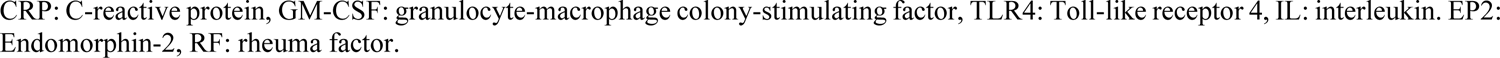
Results of multiple regression analysis with psychiatry symptom domains as dependent variables

| Dependent variables | Explanatory variables | $\beta$ | t | p | F<sub>model</sub> | df | p | R<sup>2</sup> |
| --- | --- | --- | --- | --- | --- | --- | --- | --- |
| #1. z PP | Model |  |  |  | 75.279 | 3/164 | <0.001 | 0.579 |
|  | CRP | 0.562 | 9.22 | <0.001 |  |  |  |  |
|  | GM-CSF | 0.220 | 3.77 | <0.001 |  |  |  |  |
|  | TLR4 | 0.133 | 2.38 | 0.019 |  |  |  |  |
| #2. Total BDI | Model |  |  |  | 25.844 | 3/164 | <0.001 | 0.321 |
|  | CRP | 0.374 | 4.83 | <0.001 |  |  |  |  |
|  | GM-CSF | 0.181 | 2.45 | 0.015 |  |  |  |  |
|  | TLR4 | 0.147 | 2.07 | 0.040 |  |  |  |  |
| #3. Total FF | Model |  |  |  | 83.02 | 3/164 | <0.001 | 0.603 |
|  | CRP | 0.624 | 10.96 | <0.001 |  |  |  |  |
|  | GM-CSF | 0.179 | 3.11 | 0.002 |  |  |  |  |
|  | EP2 | 0.121 | 2.29 | 0.023 |  |  |  |  |
| #4. Total HAMA | Model |  |  |  | 46.19 | 5/162 | <0.001 | 0.588 |
|  | CRP | 0.416 | 5.75 | <0.001 |  |  |  |  |
|  | GM-CSF | 0.179 | 3.00 | 0.003 |  |  |  |  |
|  | RF | 0.172 | 2.64 | 0.009 |  |  |  |  |
|  | Anti-CCP | 0.129 | 2.25 | 0.026 |  |  |  |  |
|  | TLR4 | 0.116 | 2.06 | 0.041 |  |  |  |  |
| #5. Key_BDI | Model |  |  |  | 32.31 | 2/165 | <0.001 | 0.281 |
|  | CRP | 0.427 | 5.543 | 0.000 |  |  |  |  |
|  | GM-CSF | 0.183 | 2.422 | 0.016 |  |  |  |  |
| #6. Key_HAMA | Model |  |  |  | 38.03 | 3/164 | <0.001 | 0.410 |
|  | CRP | 0.425 | 5.47 | <0.001 |  |  |  |  |
|  | GM-CSF | 0.172 | 2.46 | 0.015 |  |  |  |  |
|  | RF | 0.160 | 2.12 | 0.035 |  |  |  |  |
| #7. Z PS | Model |  |  |  | 63.19 | 2/164 | <0.001 | 0.435 |
|  | CRP | 0.447 | 6.67 | <0.001 |  |  |  |  |
|  | GM-CSF | 0.315 | 4.71 | <0.001 |  |  |  |  |

BDI: Beck's Depression Inventory, FF: FibroFatigue scale, HAMA: Hamilton Anxiety Rating Scale; Key\_BDI: key depressive symptoms, Key\_HAMA: key anxiety symptoms, z PS: composite score of all physiosomatic symptoms
CRP: C-reactive protein, GM-CSF: granulocyte-macrophage colony-stimulating factor, TLR4: Toll-like receptor 4, IL: interleukin. EP2: Endomorphin-2, RF: rheuma factor.

### Correlation of psychiatric rating scale scores and the indices of clinical severity of RA

The correlations between the psychiatric rating scale scores and the severity of RA in the entire study group (n=168) and the restricted group of RA patients (n=118) are presented in **Table 6**. The total BDI, FF, HAMA scores, and z PP scores were significantly and positively correlated with the DAS28-4, CDAI, SDAI, TJCs and SJCs, PGA, and EGA.

**Table 6.** Intercorrelation matrix of psychopathology rating scale scores and the indices of clinical severity of rheumatoid arthritis.

| Parameter | Total BDI | Total FF | Total HAMA | z PP |
| --- | --- | --- | --- | --- |
| DAS28-4 | 0.637 / 0.609 | 0.782 / 0.545 | 0.779 / 0.497 | 0.838 / 0.632 |
| CDAI | 0.596 / 0.545 | 0.733 / 0.422 | 0.751 / 0.421 | 0.799 / 0.533 |
| SDAI | 0.619 / 0.592 | 0.777 / 0.537 | 0.771 / 0.484 | 0.829 / 0.617 |
| Tender joint count | 0.625 / 0.641 | 0.711 / 0.368 | 0.752 / 0.415 | 0.799 / 0.553 |
| Swollen joint count | 0.590 / 0.449 | 0.711 / 0.332 | 0.726 / 0.322 | 0.771 / 0.425 |
| PGA | 0.511 / 0.373 | 0.720 / 0.372 | 0.723 / 0.335 | 0.762 / 0.411 |
| EGA | 0.494 / 0.325 | 0.713 / 0.340 | 0.719 / 0.322 | 0.754 / 0.374 |

Nominator: correlations are computed in patients and controls combined (n=168); denominator: correlations are computed in patients only (n=118). All correlations at $p < 0.001$ .
BDI: Beck Depression Inventory, FF: Fibro-Fatigue scale, HAMA: Hamilton Anxiety Rating Scale, z PP composite score reflecting overall psychopathology, DAS28-4: Disease Activity Score by four factors including C-Reactive Protein, CDAI: Clinical Disease Activity Index, SDAI: Simple Disease Activity Index

### Prediction of the PP scale scores using biomarkers and RA severity scores

**Table 7** shows the results of multiple regression analysis with the PP rating scale scores as dependent variables and the biomarkers and severity RA indices as explanatory variables while allowing for the effects of age, sex, BMI, and smoking. Regression #1 shows that 68.1% of the z PP composite score variance was explained by the variance on TJC and SJCs, CRP, GM-CSF (all positive), and IL-10 (inversely). **Figure 2** shows the partial regression plot of the total BDI-II score on TJC (adjusted for EP2 and IL-10) and **Figure 3** the partial regression of BDI-II score on IL-10 (adjusted for EP2 and TJC). Regression #2 shows that 52.9% of the variance in the total BDI-II score was explained by IL-10 (negatively) and EP2, TJC, and SJC (all three positively). A considerable part of the variance in the total FF score (61.4%) was explained by CRP, GM-CSF, and TJC (all positively, seen in Regression #3). Regression #4 shows that 61.6% of the variance in HAMA was explained by the combined effects of CRP, GM-CSF, and TJC (all positively). Regression #5 shows that 49.6% of the variance in Key_BDI was explained by the combined effects of TJC and SJC (both positively) and IL-10 (inversely). Regression #6 shows that 42.5% of the variance in the Key_HAMA symptoms was explained by the combined effects of CRP and CDAI. The regression explained a significant part of Key_FF variance (41.3%) on CRP, GM-CSF, PGA, and EP2 (positively correlated), and IL-10 (negative) (Regression #7). Regression #8 shows that 57.9% of the variance in z PS could be explained by TJC, SJC, CRP, and EP2 (all positively), and IL-10 (inversely). A considerable part of the variance in z Fatigue (48.0%) was explained by CRP, GM-CSF, EP2, and TJC (positively), and IL-10 (negatively) (see Regression #9).

**Figure 2.**
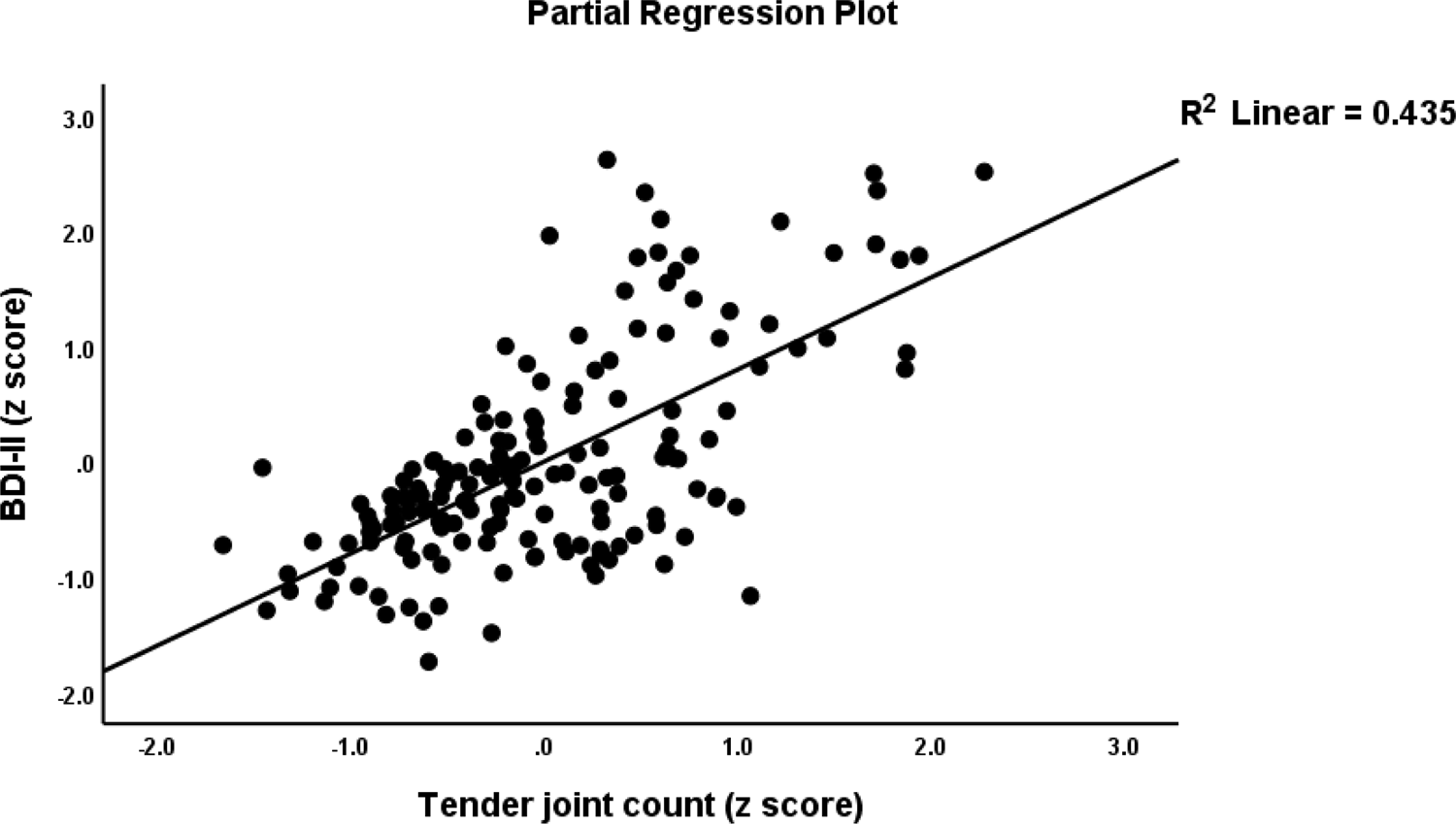
Partial regression plot of the total Depression Inventory (BDI-II) score on the tender joint count.

**Figure 3.**
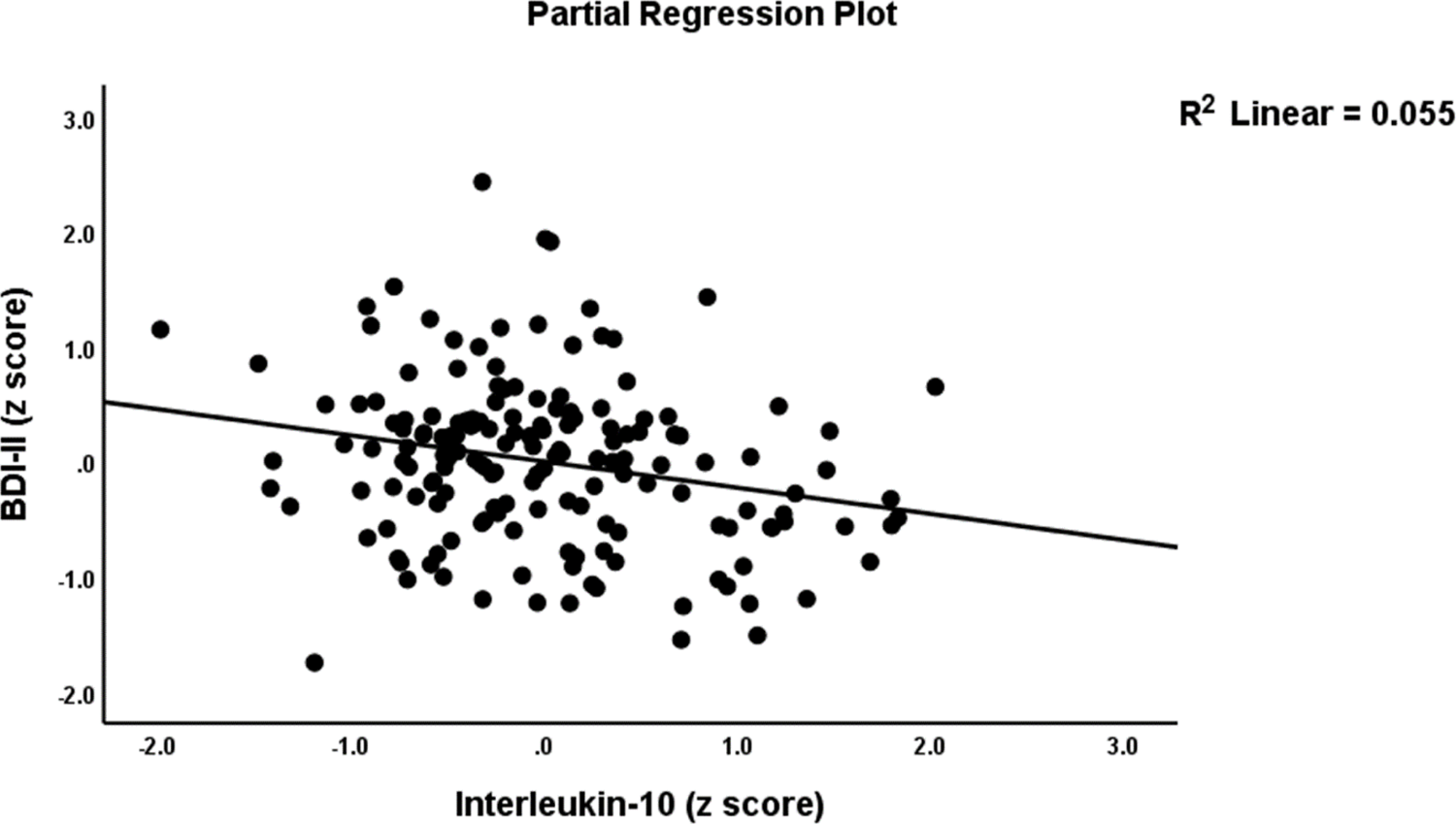
Partial regression plot of the total Depression Inventory (BDI-II) scores on interleukin-10.

**Table 7.** Results of multiple regression analysis with psychiatric symptom domains as dependent variables

| Dependent variables | Explanatory variables | $\beta$ | t | p | F<sub>model</sub> | df | p | R<sup>2</sup> |
| --- | --- | --- | --- | --- | --- | --- | --- | --- |
| #1. z PP | **Model** |  |  |  | 69.27 | 5/162 | <0.001 | 0.681 |
|  | Tender joint count | 0.445 | 5.61 | <0.001 |  |  |  |  |
|  | CRP | 0.338 | 4.94 | <0.001 |  |  |  |  |
|  | GM-CSF | 0.113 | 2.08 | 0.039 |  |  |  |  |
|  | IL-10 | -0.165 | -2.64 | 0.009 |  |  |  |  |
|  | Swollen joint count | 0.166 | 2.22 | 0.028 |  |  |  |  |
| #2. Total BDI | **Model** |  |  |  | 43.84 | 4/156 | <0.001 | 0.529 |
|  | Tender joint count | 0.673 | 7.60 | <0.001 |  |  |  |  |
|  | IL-10 | -0.267 | -3.52 | 0.001 |  |  |  |  |
|  | Swollen joint count | 0.189 | 2.17 | 0.031 |  |  |  |  |
|  | EP2 | 0.132 | 2.16 | 0.032 |  |  |  |  |
| #3. Total FF | **Model** |  |  |  | 86.83 | 3/164 | <0.001 | 0.614 |
|  | CRP | 0.508 | 7.21 | <0.001 |  |  |  |  |
|  | Tender joint count | 0.231 | 3.15 | 0.002 |  |  |  |  |
|  | GM-CSF | 0.150 | 2.56 | 0.011 |  |  |  |  |
| #4. Total HAMA | **Model** |  |  |  | 87.87 | 3/164 | <0.001 | 0.616 |
|  | Tender joint count | 0.404 | 5.52 | <0.001 |  |  |  |  |
|  | CRP | 0.353 | 5.03 | <0.001 |  |  |  |  |
|  | GM-CSF | 0.136 | 2.33 | 0.021 |  |  |  |  |
| #5. Key_BDI | **Model** |  |  |  | 53.72 | 3/164 | <0.001 | 0.496 |
|  | Tender joint count | 0.674 | 7.34 | <0.001 |  |  |  |  |
|  | IL-10 | -0.219 | -2.90 | 0.004 |  |  |  |  |
|  | Swollen joint count | 0.193 | 2.14 | 0.034 |  |  |  |  |
| #6. Key_HAMA | **Model** |  |  |  | 60.99 | 2/165 | <0.001 | 0.425 |
|  | CDAI | 0.389 | 4.25 | <0.001 |  |  |  |  |
|  | CRP | 0.305 | 3.34 | 0.001 |  |  |  |  |
| #7. Key_FF | **Model** |  |  |  | 22.63 | 5/161 | <0.001 | 0.413 |
|  | CRP | 0.327 | 3.45 | 0.001 |  |  |  |  |
|  | GM-CSF | 0.240 | 3.27 | 0.001 |  |  |  |  |
|  | PGA | 0.273 | 2.78 | 0.006 |  |  |  |  |
|  | IL-10 | -0.229 | -2.67 | 0.008 |  |  |  |  |
|  | EP2 | 0.143 | 2.11 | 0.036 |  |  |  |  |
| #8. z PS | **Model** |  |  |  | 36.64 | 6/160 | <0.001 | 0.579 |
|  | Tender joint count | 0.449 | 4.82 | <0.001 |  |  |  |  |
|  | GM-CSF | 0.181 | 2.83 | 0.005 |  |  |  |  |
|  | IL-10 | -0.305 | -4.09 | <0.001 |  |  |  |  |
|  | CRP | 0.215 | 2.72 | 0.007 |  |  |  |  |
|  | Swollen joint count | 0.188 | 2.16 | 0.032 |  |  |  |  |
|  | EP2 | 0.122 | 2.13 | 0.035 |  |  |  |  |
| #.9 z Fatigue | **Model** |  |  |  | 29.88 | 5/162 | <0.001 | 0.480 |
|  | Tender joint count | 0.404 | 4.46 | <0.001 |  |  |  |  |
|  | CRP | 0.299 | 3.49 | 0.001 |  |  |  |  |
|  | EP2 | 0.204 | 3.21 | 0.002 |  |  |  |  |
|  | IL-10 | -0.267 | -3.30 | 0.001 |  |  |  |  |
|  | GM-CSF | 0.170 | 2.43 | 0.016 |  |  |  |  |

BDI: Beck Depression Inventory, FF: Fibro Fatigue Rating Scale, HAMA: Hamilton Anxiety Rating Scale, Key\_BDI: key depressive symptoms, Key\_HAMA: key anxiety symptoms, z PP: a composite score reflecting overall psychopathology, z PS: composite score of all physiosomatic symptoms, z Fatigue: a composite score reflecting fatigue, CRP: C-reactive protein; GM-CSF: granulocyte-macrophage colony-stimulating factor; IL: interleukin; EP2: Endomorphin-2.

### PLS analysis

**Figure 4** depicts the final PLS model, which investigated the causal pathways from autoimmunity and smoking to the phenome of RA. The latter included all RA severity measures (except DAS-28 and SDAI as they comprise CRP values) as well as all key PP and PS symptoms. The RA phenome was conceived as a latent vector (reflective model) derived from these 13 clinical scores. A latent vector (in a reflective model) was extracted from IL-6, IL-10, TLR4, CRP, and GM-CSF and reflected immune activation. In addition, EM2 and KOR were combined in a single latent formative vector. All other variables, including RF, anti-CCP, smoking, FBXW7, and CD17, were entered as single indicators. With SRMR=0.048, the model’s overall fit was adequate. The phenome reflective latent vector’s construct reliability was adequate, with Cronbach =0.954, rho A=0.967, composite reliability=0.960, and average variance extracted=0.689. The loadings on this factor were greater than 0.666 at p < 0.0001 and blindfolding revealed that the construct cross-validated redundancy (0.457) was more than adequate. Furthermore, the immune activation factor construct reliabilities were adequate, with Cronbach=0.817, rho_A=0.836, composite reliability=0.872, and average variance extracted=0.578. At p<0.0001, all loadings on this factor were greater than 0.666 and blindfolding revealed that the construct cross-validated redundancy (0.276) was adequate. CTA showed that the immune activation and phenome latent vectors were not misspecified as reflective models. We discovered that 69.7 percent of the variance in the phenome latent vector was explained by immune activation, RF, anti-CCP, CD17, and MOR and that 50.2 percent of the variance in immune activation was explained by RF, anti-CCP, and smoking. Smoking and RF explained 18.1 percent of the variance in KOR+EP2. There were significant indirect effects of smoking on the phenome, mediated by immune activation (t=2.97, p=0.002) or MOR (t=1.68, p=0.046). There were also significant indirect effects of RF (t=6.82, p<0.001) and anti-CCP (t=4.59, p<0.001) on the RA phenome, which were both mediated by immune activation. All the endogenous construct indicators had positive Q^2^ Predict values, indicating that they outperformed the naive benchmark. The use of Predicted-Oriented Segmentation analysis in conjunction with Multi-Group Analysis and Measurement Invariance Assessment resulted in complete compositional invariance.

**Figure 4.**
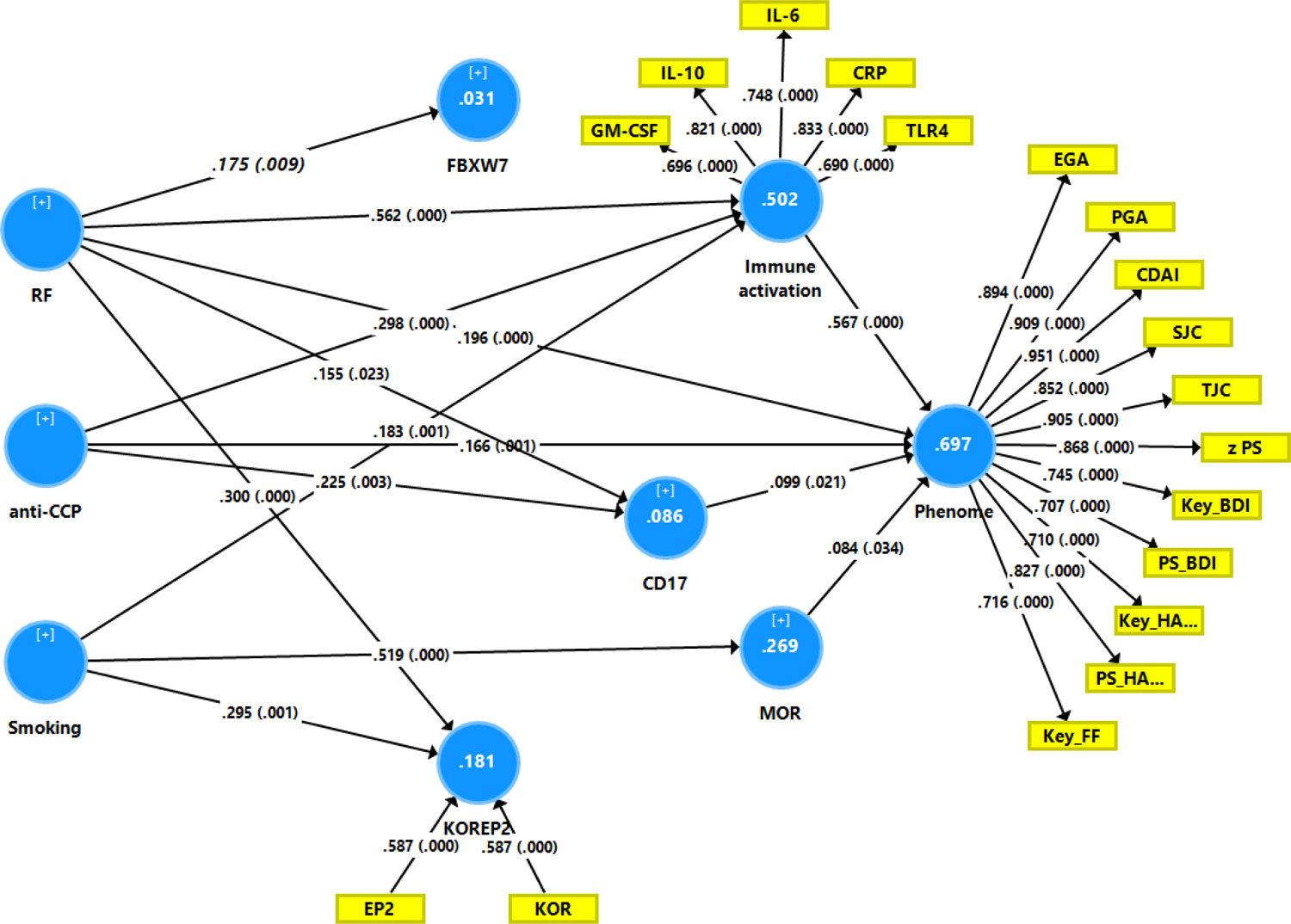
Results of Partial Least Squares PLS) path analysis. TJC: tender joint count, SJC: swollen joint count, PGA: patient global assessment, EGA: evaluator global assessment, CDAI: clinical disease activity Index, Key_FF: key symptoms of the Fibromyalgia and Chronic Fatigue Syndrome Rating (FF) scale, Key_BDI: key depressive symptoms of the Beck-Depression Inventory (BDI-II), KEY_HA: key anxiety symptoms of the Hamilton Anxiety Rating Scale (HAMA), PS_HA: physiosomatic symptoms of the HAMA, PS_BDI: physiosomatic symptoms of the BDI-II, z PS: integrated index of physiosomatic symptoms. IL: interleukin, CRP: C-reactive protein, TLR4: Toll-Like Receptor 4, GM-CSF: granulocyte-macrophage colony-stimulating factor, FBXW7: F-box/WD repeat-containing protein 7, CD17: lactosylceramide, MOR: mu-opioid receptor, KOR: kappa-opioid receptor, EP2: endomorphin-2, RF: rheumatoid factor, anti-CCP: anti-citrullinated protein antibodies.

## Discussion

### Association psychopathology and RA disease activity

The study’s first major finding is the strong relationship between depressive, anxiety, CFS, and physiosomatic severity ratings and disease activity of RA as measured with PGA, EGA, CDAI, SDAI, DAS28, TJC, and SJC. Moreover, RA patients with increased ratings on the PP rating scales also showed increased scores on all these disease activity assessments compared with RA patients without increased PP ratings. These findings extend previous papers showing that CDAI scores were significantly associated with depression ratings. ^64–66^ In another study in RA, patients with depression showed higher ratings on TJCs, EGA, PGA, DAS-28, SDAI, and CDAI. ^65^ A cross-sectional study performed in The Kenyatta National Hospital showed that depression was accompanied by increased CDAI scores and lowered quality of life. ^67^ In another study, the CDAI score was significantly associated with self-reported depression and persistent depressive symptoms. ^64^ Treatment of RA with tocilizumab, an IL-6 antagonist, may reduce depression and anxiety ratings. ^68^ Moreover, depression in RA may cause increased disabilities and reduce a positive response to treatment and the likelihood of achieving remission. ^69–72^ Singh et al (2014), reported that fatigue scores are significantly and positively associated with the DAS28, CDIA, TJC, SJC, PGA, and EGA. ^73^ Holten et al. (2021) found that a high number of RA patients suffer from fatigue (69%) and that this high prevalence decreased during treatment in association with PGA and SJC. ^74^ Many people with RA (up to 43%) show a somatization comorbidity phenotype (the PS symptoms measured in our study) and the response to treatment may be reduced in this group of patients. ^75^ All in all, these results show that RA disease activity is strongly associated with depression, anxiety, fatigue and physiosomatic symptom severity. Furthermore, there is some evidence that the relationships between depression and RA may be bidirectional because depression is also associated with an increased risk of RA and a more detrimental course of RA.^76^

It should be stressed that we included only patients with depressive, anxiety and FF symptoms due to RA and excluded patients with a lifetime history of affective and anxiety disorders (and another axis I diagnoses), CFS or Myalgic Encephalomyelitis and, therefore, our results show that RA is associated with increased severity of “secondary” depression, anxiety and fatigue. Generally, medical patients with comorbid depression have more severe symptoms and higher medical costs than those who are not depressed due to their medical condition.^77^ Future research should examine whether depression, anxiety, and CFS due to RA may affect the outcome of RA in terms of morbidity and mortality, including secondary cardio-vascular disorders, which are strongly associated with depression and CFS. ^78, 79^

Most importantly, in the current study, we found that one common factor could be extracted from the diverse PP ratings and all RA disease activity indices and that this latent vector showed excellent psychometric properties, indicating the three PP and RA disease activity scores are reflective manifestations of the same underlying construct or common core, which is the cause of the correlations among the PP and RA disease scores. It follows that increased PP ratings reflect the severity of RA and thus share the same pathophysiology as RA disease activity. Such findings exclude psychological theories that secondary PP symptoms should be ascribed to beliefs about the illness, the personal meaning attributed to stressors that accompany the illness, negative cognitions, or other folk psychology explanations.^80^ Moreover, a large part (69.7%) of the variance in this common phenome core was explained by activated immune-inflammatory pathways, autoimmunity, CD17, and changes in the EOS system. In fact, the large effects sizes show that those signaling pathways are an essential part of the phenome of depression, anxiety, and FF due to RA patients. This will now be discussed in the next subsections.

#### Immune-inflammatory pathways

The second major finding of this study is that different immune-inflammatory pathways were significantly associated with the common core and with the separate depression, anxiety, CFS, and physiosomatic scores. Previously, it was hypothesized that activated immune-inflammatory pathways may partly explain the occurrence of depression in RA. ^77^ Other authors also provided evidence that the bidirectional association between RA and depression may be explained by inflammation. ^76^ A recent review provided evidence that the relation between RA and depression can be explained by the effects of innate immune and molecular responses to inflammation. ^81^ Increased levels of proinflammatory cytokines, chemokines, type 1 interferons are the major culprits that may cause depression or explain the increased occurrence of depression in RA. ^81^

The present study showed that increased IL-6 and CRP in acute RA were significantly associated with PP symptoms due to RA. In a multivariate analysis study on RA severity, elevated serum IL-6 and CRP levels were associated with depression severity. ^44^ Moreover, patients with RA are at increased risk of developing depression, particularly if their disease activity scores and serum IL-6 levels are increased. ^44^ Increased levels of IL-6 and CRP are not only associated with major depression, ^8, 82^ but also with RA, and in the latter illness, IL-6 is associated with increased CDAI, DAS28, and bone erosions. ^83^ The severity of CFS-like symptoms in schizophrenia, another neuroimmune disorder, is associated with immune-inflammatory markers, including increased plasma IL-6. ^84^ Other authors suggested that higher levels of IL-6 are linked to both fatigue and pain and that this connection may contribute to the occurrence of depression. ^85^ Despite that, our findings show that affective symptoms and CFS belong to the same core common to both PP and RA disease activity. This phenome core is highly associated with IL-6 and CRP, suggesting that all clinical manifestations share the same immune pathophysiology. Our research results imply that the idea that depression in RA is due to pain and fatigue should be rejected.

The current study showed that increased GM-CSF, another cytokine that plays a key role in the immune-inflammatory response, is associated with PP symptoms due to RA. The GM-CSF levels are significantly increased in RA compared to healthy controls, ^86^ while GM-CSF is also overexpressed in patients with depression. ^87^ Increased TLR4 mRNA and protein expression in PBMCs and synovial tissues are observed in RA patients. ^88^ Increased TLR4 signaling and mRNA TLR4 expression are both associated with depression. ^89, 90^ Amaya-Amaya et al. suggested that interactions of pathogen-associated molecular patterns (bacterial or viral) with the TLR4 complex may be an initial inciting event in RA, ^1^ which may subsequently cause activation of the innate immune system with increased levels of proinflammatory cytokines and reactive oxygen species. ^89^ In fact, the same mechanism has been proposed to initiate major depression and CFS. ^91, 92^

In our study, increased levels of IL-10 were detected in RA, and they were positively associated with increased RA disease activity and PP assessments. In RA, IL-10 levels are positively correlated with RF, anti-CCP, and CRP, ^15, 93^ while there is also a well-established link between depression and elevated IL-10. ^39, 52, 94^ The severity of CFS-like symptoms in schizophrenia is associated not only with increased IL-6 but also IL-10 levels. ^84^ Interestingly, we detected that after considering the effects of inflammatory mediators (TJC, CRP, and GM-CSF) in regression analyses, IL-10 was inversely associated with depression, fatigue, physiosomatic, and overall PP ratings. These findings suggest that immune-inflammatory responses coupled with relative decrements in IL-10 may cause increased PP burden in RA. IL-10 has been shown to inhibit IL-6, TNF-α, and GM-CSF production from immune cells ^95^ and enhance B cell differentiation to cells secreting IgG, IgM, and IgA, ^96^ resulting in increased RF and IgG-RF production by B cells in peripheral blood. Moreover, IL-10 is localized in the synovial membrane lining layer, the site of monocyte migration, and inhibits proinflammatory cytokines in RA. ^97, 98^ In depression, IL-10 is one important component of the “compensatory immune-regulatory system” (CIRS) which tends to down-regulate the primary immune-inflammatory response. ^39, 99^

Maes et al. ^77^ reviewed that activated immune-inflammatory pathways and autoimmune and nitro-oxidative pathways may explain the occurrence of depression due to RA and other peripheral immune, autoimmune and neuroinflammatory disorders and that the bidirectional associations between affective disorders and RA may be explained by increased neuro-immune and neuro-oxidative signaling. Indeed, depression is accompanied by autoimmune responses to oxidative-specific neoepitopes (including malondialdehyde and azelaic acid) and nitroso adducts. ^36^ There is now evidence that O&NS pathways play a key role in RA, for example, by causing damage in the joints and inducing apoptotic processes in rheumatoid synovium and articular cartilage. ^100, 101^ In RA as well as in depression, there are multiple multidirectional interconnections among immune-inflammatory and O&NS pathways leading to a vicious circle between these two pathways. Moreover, such O&NS pathways also play a role in CFS and anxiety disorders, ^102, 103^ while similar autoimmune responses are also observed in depression and CFS. ^43^

#### Other biomarkers of PP due to RA

The current study found that PP symptoms were also associated with diverse alterations in the EOS and CD17 but only marginally with FBXW7. However, it should be stressed that the impact of these three biomarkers was modest compared to the major effect of immune-inflammatory pathways. Major depression is associated with elevated levels of MOR and β-endorphins, ^26^ and the synovial membrane probably produces β-endorphin. ^104^ In recent years, substantial research points to the role of the EOS and their receptors in response to stress, mood regulation, and the pathophysiology of MDD. ^52^ Activation of immune-inflammatory pathways is likely responsible for circulating levels of β-endorphin and other EOS markers (such as MOR and KOR) during inflammatory conditions. ^105, 106^ Successive expression of chemokines and adhesion molecules, primarily CXCR2 ligands, L and P selectins, α4/β2 integrins, and intercellular adhesion molecule 1 orchestrate the recruitment of opioid-containing leukocytes from the circulation to the site of inflammation. ^107–109^ EOS peptides and receptors are secreted during immune activation, and most of them act as CIRS compounds. ^26, 52, 105^

In the current study, we observed that lactosylceramide levels significantly increased in RA compared to controls and were marginally associated with the common phenome core extracted from the PP and RA disease activity data. Some ceramides, including CD17, are significantly elevated in depressed patients. ^48–50^ This is important because lactosylceramide plays a role in anxiety, neuroinflammation, reduced neurogenesis and neuronal survival, and neurodegenerative processes. ^110–112^ Moreover, inflammatory mediators, including TNF-α, activate the synthesis of lactosylceramide, which in turn activates ROS and α-type cytosolic phospholipase A2 (cPLA2α), which releases arachidonic acid, another inflammatory mediator. ^113^ Furthermore, lactosylceramide activates NADPH oxidase and iNOS and increases the production of superoxide radicals NO. ^22, 23^ As such, the increased levels of lactosylceramide may participate in the immune-inflammatory pathophysiology of RA and the onset of depressive symptoms in RA. Moreover, in an antigen-induced RA model, acid sphingomyelinase, the lysosomal enzyme which hydrolyses membrane sphingomyelin to ceramide, appears to be associated with joint swelling and increased levels of proinflammatory cytokines. ^114^ The lactosylceramide signaling pathways cause oxidative stress, inflammation, mitochondrial dysfunctions, and atherosclerosis ^115^ and, therefore, could play a role in RA-associated cardio-vascular disease.

In contrast to the a priori hypothesis, we did not find lowered levels of FBXW7 in PP due to RA. Such changes could have contributed to the onset of depressive symptoms in RA. On the contrary, the FBXW7 levels showed very marginal positive associations with the PP scores.

## Conclusions

Depression, anxiety, and CFS-like symptoms due to RA are reflective manifestations of the phenome of RA and are mediated by the effects of the same immune-inflammatory, autoimmune, EOS, and lactosylceramide pathways that underpin the pathophysiology of RA.

## Data Availability

The dataset generated during and/or analyzed during the current study will be available from MM upon reasonable request and once the authors have fully exploited the dataset.

## Competing interests

The authors declare that they have no known competing financial interests or personal relationships that could have influenced the work reported in this paper.

## Funding

There was no specific funding for this specific study.

## Author’s contributions

All authors contributed to the writing up of the paper. MM performed statistical analyses.

All authors revised and approved the final draft.

## Acknowledgements

We thank the staff of Al-Sader Teaching Hospital in Al-Najaf province-Iraq for their assistance in the samples collection.

## Compliance with Ethical Standards

### Disclosure of potential conflicts of interest

The authors declare no conflicts of interest which are relevant to the content of this article.

### Research involving Human Participants and/or Animals

The ethics committee (IRB) of the University of Kufa, Iraq (243/2020), which complies with the International Guideline for Human Research Protection, approved the study as required by the Helsinki Declaration.

### Informed consent

Before taking part in the study, all participants provided written informed consent.

## Notes

### Competing Interest Statement

The authors have declared no competing interest.

